# Machine Learning Methods to Predict Survival in Patients Following Traumatic Aortic Injury

**DOI:** 10.1101/2021.07.28.21261166

**Authors:** Nisreen Shiban, Joshua Gaul, Henry Zhan, Andrew Elhabr, Nima Kokabi, Jamlik-Omari Johnson, Tarek Hanna, Justin Schrager, Judy Gichoya, Imon Banerjee, Hari Trivedi

**Affiliations:** Emory University, Atlanta, GA; Georgia Institute of Technology, Atlanta, GA

## Abstract

The National Trauma Data Bank (NTDB) is a resource of diagnostic, treatment, and outcomes information in trauma patients. We leverage the NTDB and machine learning techniques to predict survival following traumatic aortic injury. We create two predictive models using the NTDB – 1) using all data and, 2) using only data available in the first hour after arrival (prospective data). Seven discriminative model types were tested before and after feature engineering to reduce dimensionality. The top performing model was XGBoost, achieving an overall accuracy of 0.893 using all data and 0.855 using prospective data. Feature engineering improved performance of all models. Glasgow Coma Scale score was the most important factor for survival, and thoracic endovascular aortic repair was more common in patients that survived. Smoking, pneumonia, and urinary tract infection predicted poor survival. We also note concerning disparities in outcomes for black and uninsured patients that may reflect differences in care.

## Introduction

Traumatic aortic injury (TAI) is the second leading cause of death in multi-trauma patients and requires urgent management ^1^. Although only 38% of patients survive following acute aortic injury, better management at the scene of the accident and quicker transportation to hospitals has led to improvements in overall survival, with only 4% of patients dying en route ^2^. Nonetheless, mortality around the time of the injury remains high, estimated at 20% in the 24 hours following hospital admission ^3^.

Retrospective studies around predicting mortality in TAI show that thoracic endovascular aortic repair (TEVAR) is associated with improved outcomes compared with open repair, specifically lower mortality and lower incidence of spinal cord ischemia ^4,5^. These works predominantly report the effects of non-operative management, TEVAR, or open aortic repair and control only for major demographic factors when evaluating outcomes. However, there are no comprehensive studies that include more detailed comorbidities, complications, and injury types that may affect outcomes in patients with TAI.

Modern machine learning (ML) methods and advances in hardware have enabled development of robust prediction models that integrate thousands of features, far beyond what is possible in traditional multivariate analysis. In Emergency Medicine, ML models have been developed for a variety of pathologies, including acute kidney injury, influenza, and sepsis ^6–8^. In this study, we aimed to develop an ML model to predict survival in TAI using detailed patient information available on arrival and throughout the hospital stay.

To gather sufficient data for evaluation of outcomes in TAI, we leveraged the National Trauma Data Bank (NTDB) - a large data repository that encompasses a wide variety of traumatic injuries, interventions, and outcomes in trauma patients^9^. The NTDB is compiled annually by the American College of Surgeons (ACS) using standardized data contributions from trauma hospitals across the U.S. Several studies have applied machine learning techniques to the NTDB to investigate clinical problems such as traumatic brain injury (TBI) and overall trauma severity, however these studies often focus only on limited portions of the available data or create trauma models that are too general for more rare pathologies such as TAI. For example, Abujaber *et al* discard 30% of patients with TBI due to missing data and use a hand-selected list of features rather than the entirety of data available ^10^. Gorczyca *et al* created a new general trauma severity model but do not report performance for any specific pathologies ^11^.

In this work, we leverage all available data fields in the NTDB to answer a specific clinical question and implement feature engineering techniques to improve model performance. These pre-processing techniques are generalizable regardless of the clinical application, and therefore can be used to more easily create pathology-specific models that may highlight patterns in patient outcomes. We also report the performance of multiple types of ML architectures and provide explainable results with feature importances from our top-performing model.

## Methods

### Cohort Selection

We focused on NTDB data from 2011 to 2015 for uniformity in international classification of disease 9^th^ edition (ICD-9) diagnostic codes. There were 5.1M recorded trauma incidents with each row in the dataset representing one incident. Individual patients are not tracked in the dataset so multiple incidents per patient could not be evaluated.

### Patient Selection and Outcomes Categorization

To identify incidents with TAI, we filtered based on ICD-9 diagnostic codes of 901.0, 902.0, and 441.* which are specific to dissection or other injury of the thoracic or abdominal aorta, yielding 12,435 unique incidents. The NTDB contains 18 possible patient dispositions ranging from discharged, deceased, or transfer to various facilities. Outcomes were binarized as survival (alive) or non-survival (deceased) based on these codes (Figure 1). Specifically, dispositions that included deceased, expired, or hospice were considered as the deceased class, and all other codes including discharge or transfer to another short term or long-term care facility were considered as the alive class. Patients who did not have disposition information were excluded, yielding 9,294 remaining incidents.

**Figure 1.**
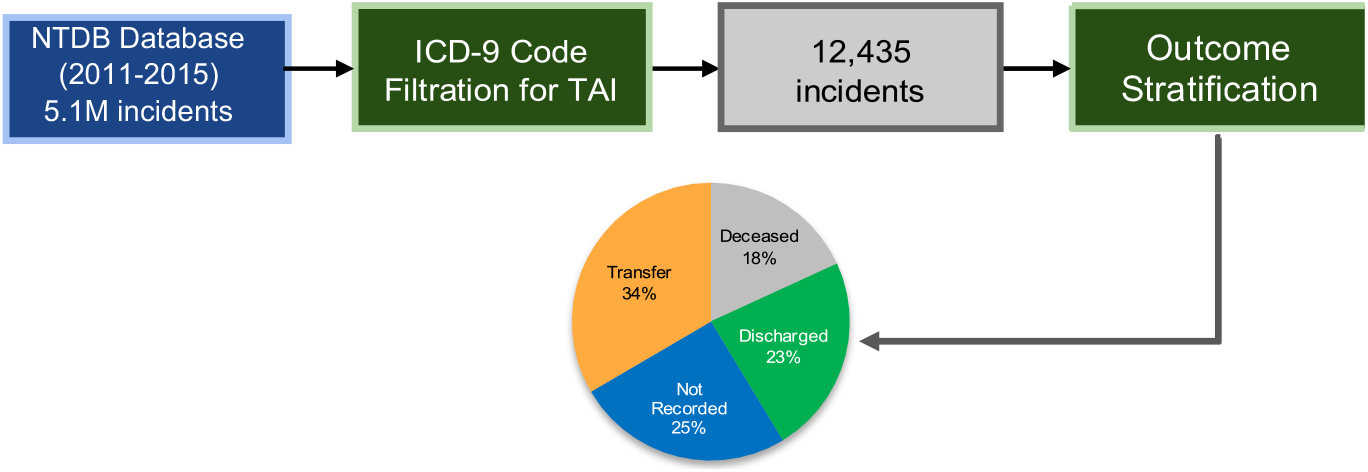
Schematic of patient selection. Traumatic aortic injury was identified using ICD-9 codes. Outcomes were identified using the *disposition* field in the NTDB. Blue represent the deceased class, yellow and orange represent the alive class, and gray represents cases that were excluded due to lack of disposition information.

### Data Elements and Model Types

There are over 23,000 discrete data elements in the NTDB, including ICD-9 diagnostic (DCODE) and procedural (PCODE) codes, abbreviated injury scale (AIS) codes, emergency medical service (EMS) response time, emergency codes (ECODE), method of arrival, ED vital signs, patient demographics, and payment methods.

Because the NTDB does not include timestamps for most data (with the exception of PCODEs), we developed two models using different data elements: **1) *Full Data* model - using all available data throughout the hospitalization, and 2) *Prospective* model – including only data available within the first hour of presentation, thereby excluding DCODEs, complications, and any PCODEs after the first hour**.

### Feature Engineering

Each data element is contained within individual .csv files that are linked by trauma incident keys. Data was imported into Pandas ^12^ dataframes inside of Python ^13^. Empty fields for each feature were replaced with a negative value to avoid the need to removal the entire sample while still allowing the model to effectively ignore the feature for that sample. To make the data more manageable for machine learning models, we employed several feature engineering techniques on each data element that are detailed below. These techniques can be employed for many different machine learning projects for the NTDB, although they may have to be tailored based on the patient cohort and clinical problem of interest. An overview of feature engineering is presented in Figure 2 and details on each major data type are outlined subsequently.

**Figure 2.**
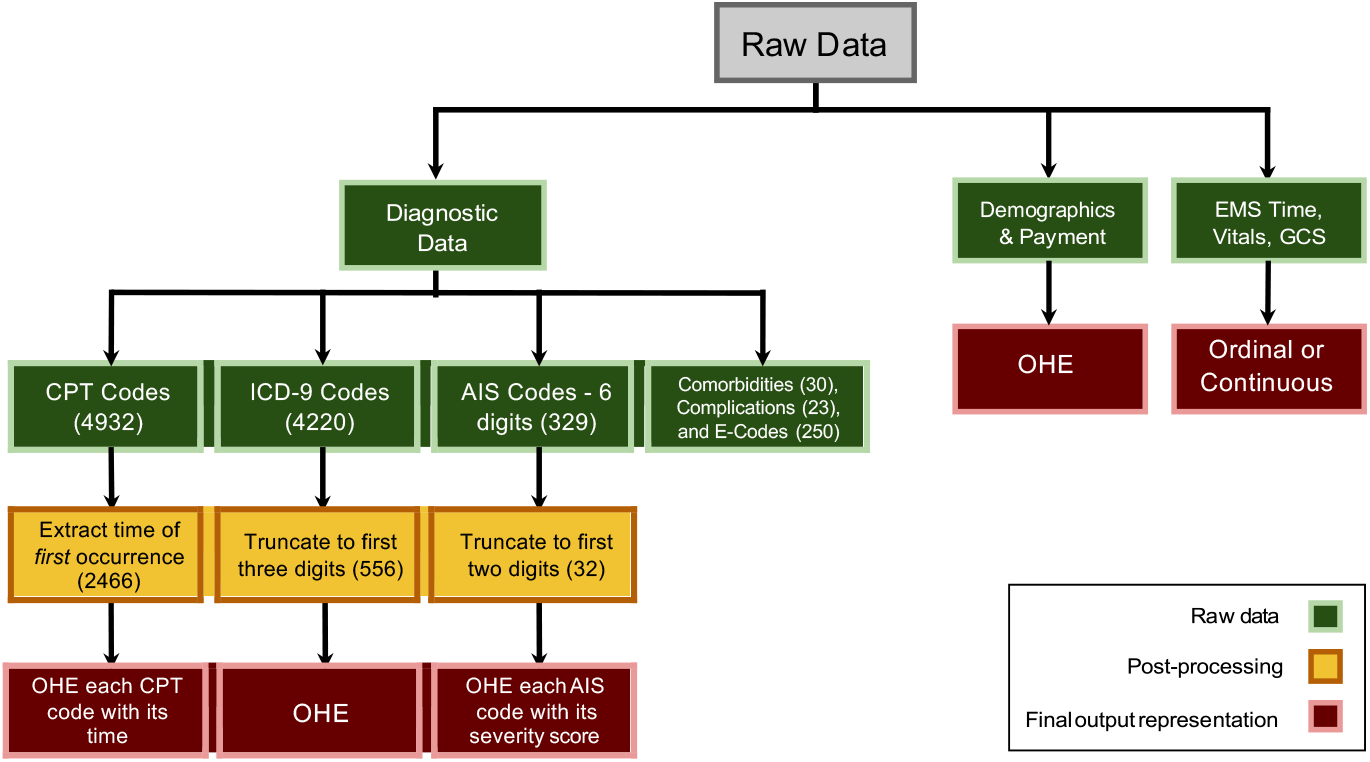
Overall schematic of feature engineering to reduce data sparseness and improve model performance. Raw data is represented in green, post-processing steps to reduce dimensionality are in yellow, and final data representation is in red. OHE = one hot encoding.

#### Diagnostic Codes (DCODE)

The ICD-9 database contains over 13,000 unique possible which and are represented for each incident using the DCODE variable. Coding using ICD-10 began in 2016 so these cases were excluded. ICD-9 codes typically consist of a stem 3 digits indicating a major category, followed by a decimal and 2 more digits indicating finer details. Our dataset contained 4220 unique DCODEs, and original attempts to train a model with all 4220 codes were unsuccessful due to data sparsity. To reduce dimensionality while maintaining major diagnostic categories, we truncated the final two digits and only considered the parent class in ICD-9 terminology, resulting in 556 remaining unique DCODEs. For instance, the original DCODE 850.12 denotes “Concussion, with loss of consciousness from 31 to 59 minutes.” When truncated to 850, the parent class of “Concussion” remains, retaining valuable clinical information.

#### Procedure Codes (PCODE)

ICD-9 procedural codes are represented by the PCODE variable. Each trauma incident contains one list of PCODEs and a second list representing the time of occurrence of each PCODE. The original size of PCODEs variable for our dataset was 4,932 unique values. Similar to DCODEs, representing the raw data created a very sparse dataset. Initially, we removed the final two digits from the PCODE to reduce dimensionality, but this removed valuable procedure details. We also attempted encoding the PCODE vector using an autoencoder model, but this eliminated the time of occurrence from the data. Ultimately, the best results were obtained by retaining each PCODE in its original form and encoding the time of *first* occurrence for each PCODE after one hot encoding. For example, for a given incident with original PCODE list of [793.19, 39.73, and 793.13] with times of [30, 90, 360] minutes, the final data would be represented as only two elements: [793.19, 30] and [39.73, 90] with removal of the second occurrence of 793.13.

#### Abbreviated Injury Scale Score

The Abbreviated Injury Scale (AIS) Score is represented by a PREDOT code and severity score. In the PREDOT code, the first digit denotes the body region of the injury (head, face, neck, thorax, etc.), the second digit denotes the type of anatomic structure (vessels, nerves, organs, skeletal, etc.), and digits 3-6 describe the nature and level of the injury (abrasion, contusion, amputation, burn, etc.). The original dataset contained 329 unique AIS codes with a sparse feature space. To reduce dimensionality, we retained only the first two digits of the PREDOT code resulting in 32 remaining features.

#### Other Features Included in Model

Vital signs include pulse, systolic blood pressure (SBP), Glasgow Coma Scale Total (GCSTOT), and oxygen saturation (OXYSAT) on arrival. The GCS total is represented by integers 1-15. Pulse, SBP, and OXYSAT are represented as continuous variables. EMSMINS represents the elapsed time between EMS transport dispatch to its arrival on the scene.

COMORBID represents pre-existing comorbidities for the patient and is represented by 25 unique elements including diabetes, hypertension, obesity, etc. Three elements - “other”, “not applicable” and “not recorded”, were removed.

COMPLIC represents any complications that occurred after patient arrival, and is represented by 25 unique elements such as pulmonary embolism, pneumonia, or surgical site infections. Similar to comorbidities, we removed the elements “other”, “not applicable” and “not recorded.”

ECODE represents the ICD-9 external cause of injury, such as motor vehicle, fall, struck by/against, firearm, or poisoning. There were 250 ECODEs in our dataset, and no feature engineering was performed.

Finally, demographic information includes age, gender, race, and ethnicity. Payment information includes the method of payment from the patient and includes elements such as self-pay, Medicare, government, or private insurance. Demographics and payment information were included in their original forms.

**Table 1.** Results of feature engineering, netting a 65% decrease in total features for the dataset. This allows models to converge more easily, resulting in better model performance. Feature reduction also increased model interpretability and generalizability by decreasing the chance of overfitting.

| Data Type | Original Size | Final Size | % Reduction |
| --- | --- | --- | --- |
| Comorbidities | 30 | 30 | 0 |
| Pulse | 1 | 1 | 0 |
| Systolic Blood Pressure | 1 | 1 | 0 |
| GCS Total | 1 | 1 | 0 |
| Oxygen Saturation | 1 | 1 | 0 |
| E-codes | 250 | 250 | 0 |
| Age | 1 | 1 | 0 |
| Gender | 3 | 3 | 0 |
| Race | 8 | 8 | 0 |
| Ethnicity | 4 | 4 | 0 |
| Facility Key | 1 | 1 | 0 |
| D-codes | 4220 | 556 | 86.82 |
| Complications | 23 | 23 | 0 |
| Hospital Discharge | 18 | 2 | 88.89 |
| Payment | 12 | 12 | 0 |
| AIS Codes | 329 | 32 | 90.27 |
| EMSMINS | 1 | 1 | 0 |
| P-codes and Time | 4932 | 2466 | 50 |
| All features | 9907 | 3444 | 65.24 |

### Discriminative Model Selection and Evaluation

Multiple classification models were tested, including Logistic Regression (LR), k-Nearest Neighbors (KNN), extreme gradient boosting (XGBoost), radial basis function SVM (RBF SVM), Multilayer Perceptron (MLP), Random Forest (RF) ^14^, and an ensemble of these models. Oversampling was used to handle the class imbalance between alive and deceased for all models. Each model was trained using five-fold cross validation and grid search over the hyperparameters. Using the parameters that maximize recall on the validation set, the model predicts and outputs the labels for the test set. For each model, the class-wise accuracy, recall, precision, and F1-score are reported.

## Results

### Patient Summary Statistics

Of 9,294 trauma incidents, 73.8% (6,855) occurred in males and 26.2% (2,437) occurred in females and two were of unknown gender. Survival was 74.3% in males and 79.8% in females. Age was similar between groups with mean age of 42.2 +/- 30.0 years in the alive class and 39.7 +/- 33.1 years in the deceased class (Table 1, Figure 8). Overall racial distribution of patients was 66.3% white and 17.5% black, however death rate was higher in blacks (37.9%) as compared to whites (19.7%). Sample size for other races was too small to draw conclusions. There were also disparities in outcomes based on payment method, with death rates between 14.9 - 17.1% for privately insured patients, as compared to death rates of 20.5 – 41.7% for Medicare, Medicaid, and self-pay patients.

**Table 1.**
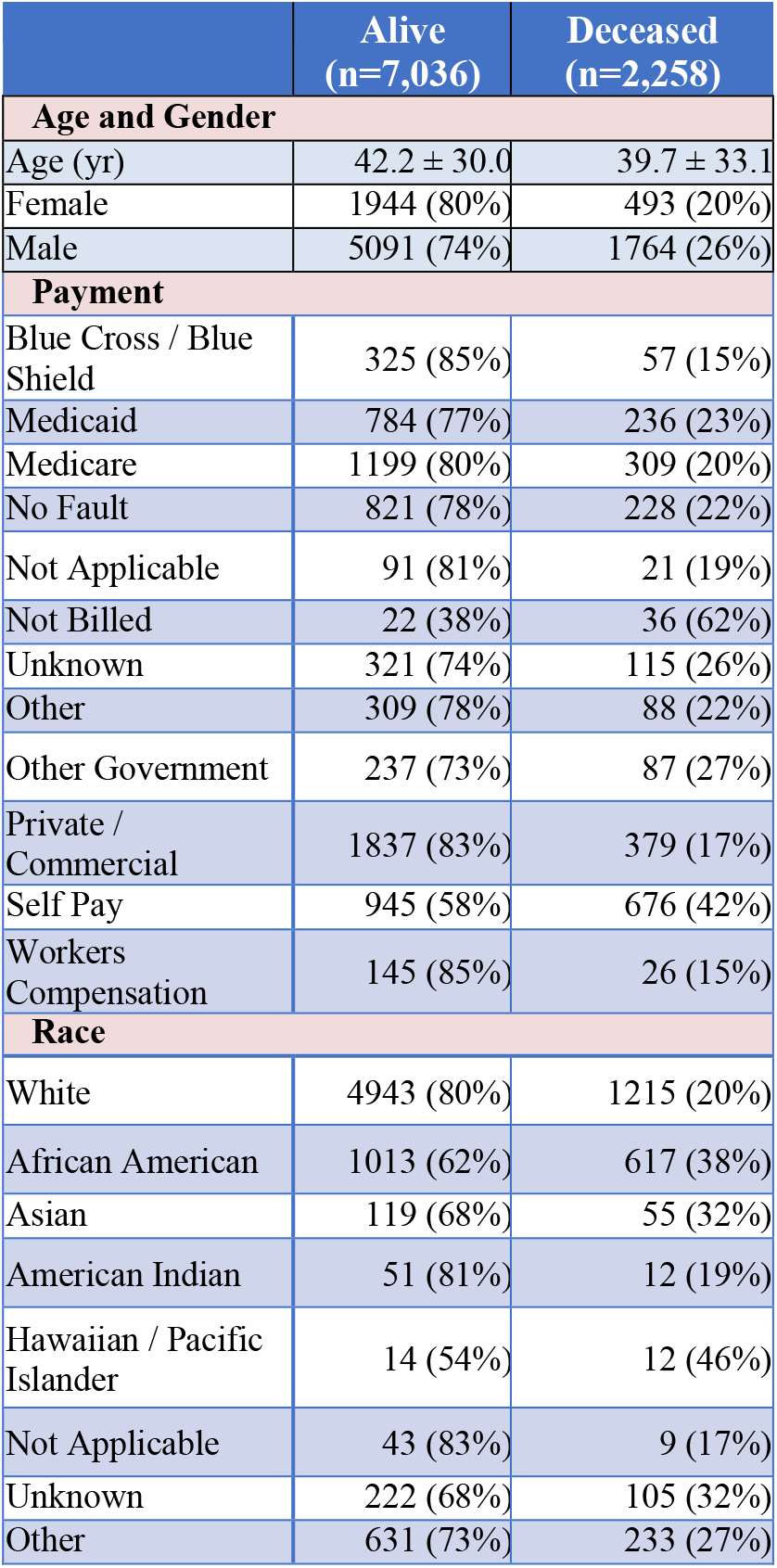
Demographics distribution of patients. Overall survival was similar between genders, although there were significantly more male patients than female overall. Self-pay, Medicare, Medicaid, and other government coverage patients overall had higher rates of death as compared to privately insured patients. African American patients also had higher rates of death than white patients.

### Model Performance

Feature engineering netted a 65.2% compression in the feature space, from an initial size of 9,907 to a final total of 3,444 features. DCODEs, PCODEs, and AIS codes were responsible for the greatest reduction (Table 2).

Overall, the *Full Data* model performed slightly better than the *Prospective* model and both models performed better after feature engineering, although absolute performance differences varied based upon the model architecture. Performances was averaged over 100 iterations of random bootstrapping and results are summarized in Table 3,

**Table 3.**
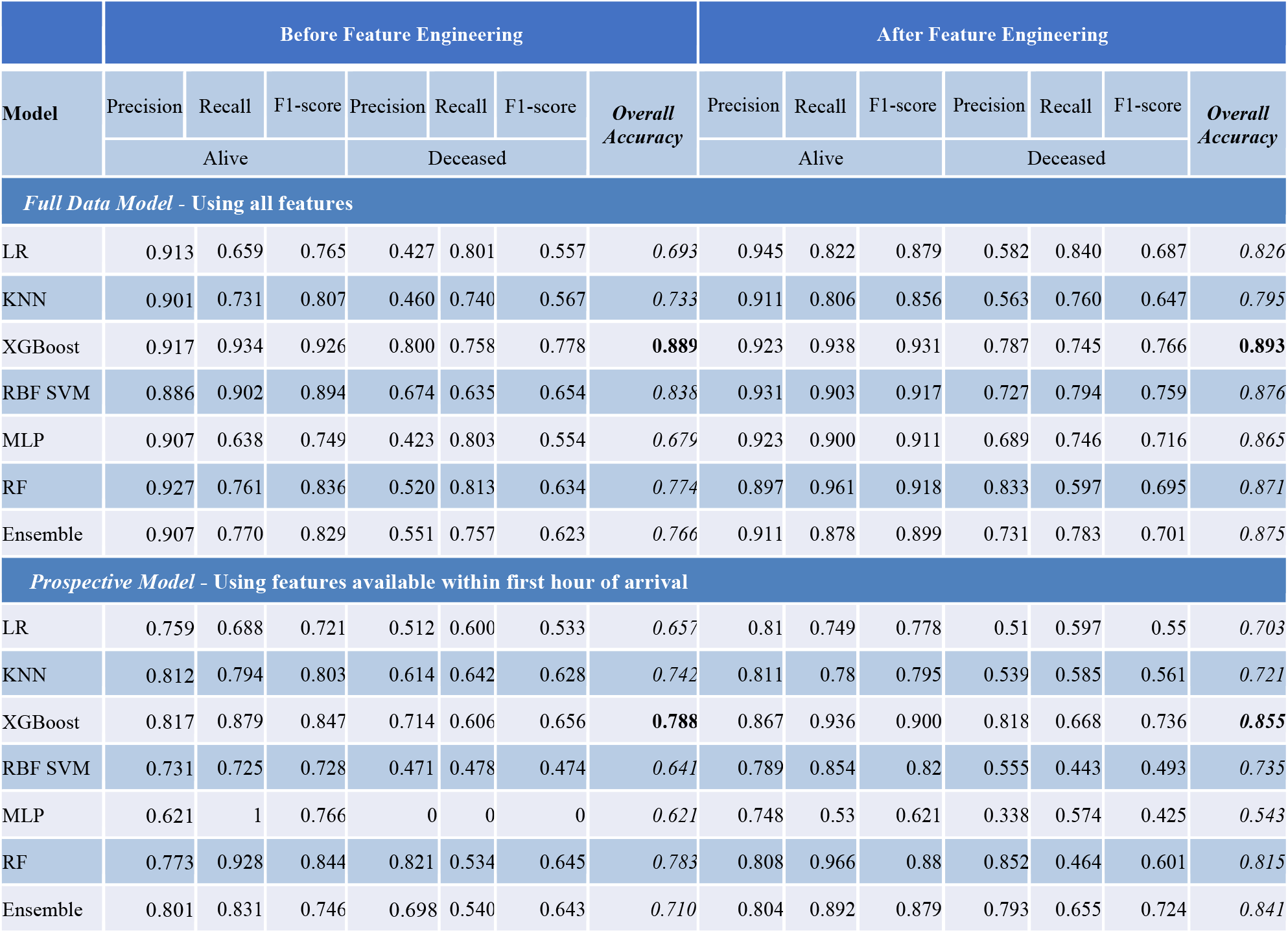
Model performances before and after feature engineering for the *Full Data* and *Prospective* models. Feature engineering resulted in increases in performance all models, although gains were modest for XGBoost. Performance decreased slightly overall for the *Prospective* model, with the biggest performance decrease in recall for the deceased class.

|  | Before Feature Engineering |  |  |  |  |  |  | After Feature Engineering |  |  |  |  |  |  |
| --- | --- | --- | --- | --- | --- | --- | --- | --- | --- | --- | --- | --- | --- | --- |
| Model | Precision | Recall | F1-score | Precision | Recall | F1-score | Overall Accuracy | Precision | Recall | F1-score | Precision | Recall | F1-score | Overall Accuracy |
|  | Alive |  |  | Deceased |  |  |  | Alive |  |  | Deceased |  |  |  |
| Full Data Model - Using all features |  |  |  |  |  |  |  |  |  |  |  |  |  |  |
| LR | 0.913 | 0.659 | 0.765 | 0.427 | 0.801 | 0.557 | 0.693 | 0.945 | 0.822 | 0.879 | 0.582 | 0.840 | 0.687 | 0.826 |
| KNN | 0.901 | 0.731 | 0.807 | 0.460 | 0.740 | 0.567 | 0.733 | 0.911 | 0.806 | 0.856 | 0.563 | 0.760 | 0.647 | 0.795 |
| XGBoost | 0.917 | 0.934 | 0.926 | 0.800 | 0.758 | 0.778 | 0.889 | 0.923 | 0.938 | 0.931 | 0.787 | 0.745 | 0.766 | 0.893 |
| RBF SVM | 0.886 | 0.902 | 0.894 | 0.674 | 0.635 | 0.654 | 0.838 | 0.931 | 0.903 | 0.917 | 0.727 | 0.794 | 0.759 | 0.876 |
| MLP | 0.907 | 0.638 | 0.749 | 0.423 | 0.803 | 0.554 | 0.679 | 0.923 | 0.900 | 0.911 | 0.689 | 0.746 | 0.716 | 0.865 |
| RF | 0.927 | 0.761 | 0.836 | 0.520 | 0.813 | 0.634 | 0.774 | 0.897 | 0.961 | 0.918 | 0.833 | 0.597 | 0.695 | 0.871 |
| Ensemble | 0.907 | 0.770 | 0.829 | 0.551 | 0.757 | 0.623 | 0.766 | 0.911 | 0.878 | 0.899 | 0.731 | 0.783 | 0.701 | 0.875 |
| Prospective Model - Using features available within first hour of arrival |  |  |  |  |  |  |  |  |  |  |  |  |  |  |
| LR | 0.759 | 0.688 | 0.721 | 0.512 | 0.600 | 0.533 | 0.657 | 0.81 | 0.749 | 0.778 | 0.51 | 0.597 | 0.55 | 0.703 |
| KNN | 0.812 | 0.794 | 0.803 | 0.614 | 0.642 | 0.628 | 0.742 | 0.811 | 0.78 | 0.795 | 0.539 | 0.585 | 0.561 | 0.721 |
| XGBoost | 0.817 | 0.879 | 0.847 | 0.714 | 0.606 | 0.656 | 0.788 | 0.867 | 0.936 | 0.900 | 0.818 | 0.668 | 0.736 | 0.855 |
| RBF SVM | 0.731 | 0.725 | 0.728 | 0.471 | 0.478 | 0.474 | 0.641 | 0.789 | 0.854 | 0.82 | 0.555 | 0.443 | 0.493 | 0.735 |
| MLP | 0.621 | 1 | 0.766 | 0 | 0 | 0 | 0.621 | 0.748 | 0.53 | 0.621 | 0.338 | 0.574 | 0.425 | 0.543 |
| RF | 0.773 | 0.928 | 0.844 | 0.821 | 0.534 | 0.645 | 0.783 | 0.808 | 0.966 | 0.88 | 0.852 | 0.464 | 0.601 | 0.815 |
| Ensemble | 0.801 | 0.831 | 0.746 | 0.698 | 0.540 | 0.643 | 0.710 | 0.804 | 0.892 | 0.879 | 0.793 | 0.655 | 0.724 | 0.841 |

The top performing architecture for both the *Full Data* and *Prospective* models was XGBoost using 1,000 trees, with overall accuracies of 0.893 and 0.855, respectively. Area under the ROC and precision-recall curves were only slightly decreased for the *Prospective* model compared to *Full Data* model (Figure 3). Performance suffered most for recall in the deceased class for the *Prospective* model. The underlying reason for top performance of the XGBoost model can be attributed to the way in which weak learners are converted to strong learners through weight adjustments over multiple model iterations. This reduces bias from the model and improves accuracy. Other advantages of XGBoost are that it is highly scalable/parallelizable and has high execution efficiency.

**Figure 3.**
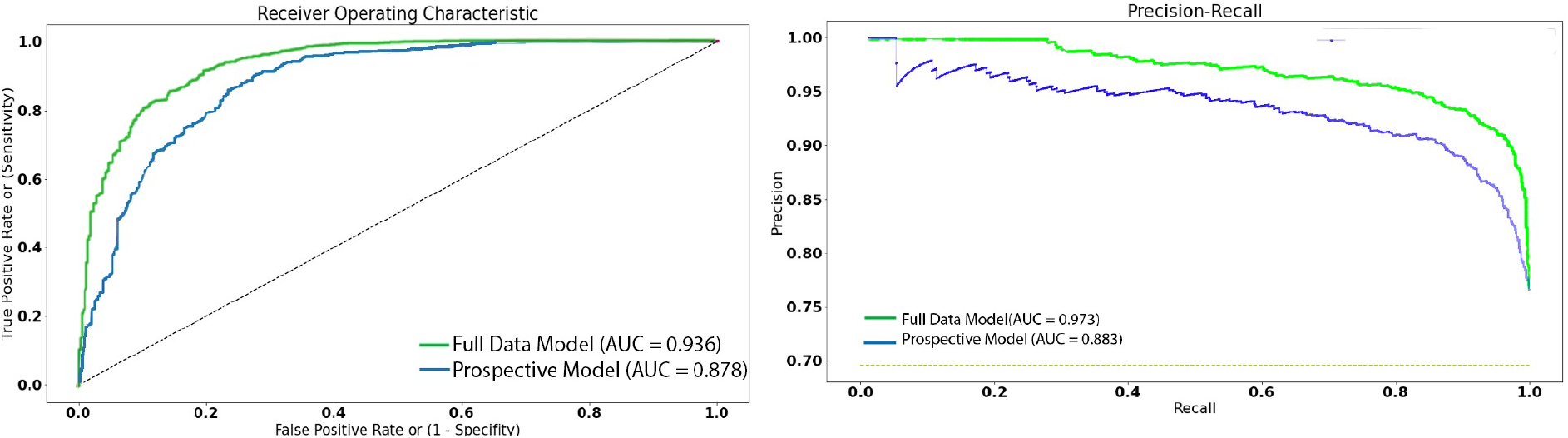
Receiving operating characteristic (left) and precision-recall curve (right) for the *Full Data (green)* and *Prospective (blue)* models. There was a small drop in all metrics for the *Prospective* model (blue) which excluded DCODES, complications, and any PCODE after the first hour. However both models still demonstrate high predictive performance.

### Feature Importances

#### Overall Top Features

Top feature importances for the both the *Full Data* and *Prospective* models are shown in Figure 4, demonstrating that the Glasgow Coma Scale (GCS) score is the most important feature predicting survival. This, along with feature 4 (AIS for head injury) and feature 6 (ICD-9 for concussion) in the *Full Data* model, suggests that patients with concurrent head trauma are much less likely to survive, either due to direct neurologic damage or possibly from autonomic dysregulation. A cohort study conducted by Indiana University School of Medicine and Rehabilitation Hospital showed that the hazard of death after traumatic brain injury (TBI) was higher for all TBI injury classification categories as compared to non-head traumatic injuries during the entire follow-up period ^15^. For the *Prospective* model, presence of early thoracotomy was a predictor for poor survival as was being a self-pay patient. The latter could be related to multiple confounding factors, including self-pay patients being more likely to be in previously poor health, indigent, or more severely injured and presenting to inner-city hospitals. Smoking was an important comorbidity for both models, likely related to underlying vascular disease. Complications of pneumonia and urinary tract infection important predictors considered only in the *Full Data* model since they occur after hospitalization. Lack of insurance and advanced directives limiting care were both important contributors to death.

**Figure 4.**
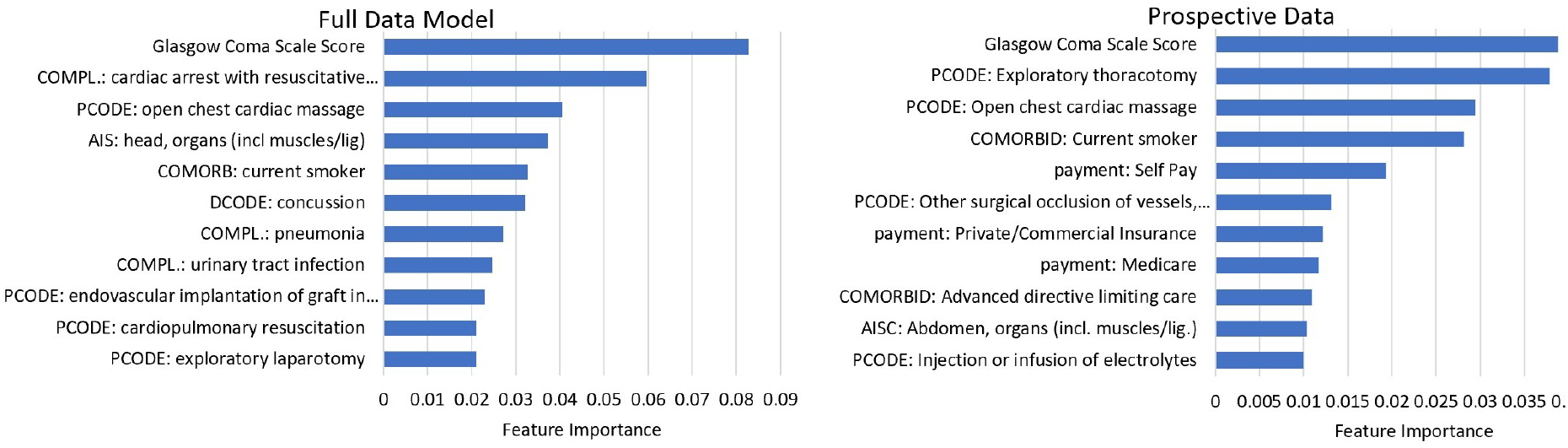
Overall feature importance for all data in the *Full Data* (left) and *Prospective Data* (right) models. Glasgow Coma Scale score was the most important feature for both models, indicating that concomitant head injury is a strong predictor of survival. Smoking history was highly predictive for both models, and complications of pneumonia and urinary tract infection after hospitalization was predictive in the *Full Data* model. Procedures including open thoracotomy within the first hour, cardiac massage, and resuscitation were also associated with poor survival.

#### Procedure Codes

Endovascular graft implantation was the third most important procedure code in the *Full Data model* indicating that this is common in patients that survive (Figure 5). This is corroborated by previous studies highlighting improved outcomes in patients undergoing TEVAR, and that patients presenting to locations where this is unavailable may have worse outcomes. Results from multicenter clinical trials have demonstrated many early benefits of TEVAR as compared with standard surgical repair, such as less blood loss and transfusion requirement, reduced ICU utilization, shorter procedure times, decreased length of hospital stay, lower rates of major adverse events, and quicker recovery^16^. The mean time for each of the top procedures was calculated between the groups, and there was no statistical difference, although both exploratory thoracotomy and laparotomy trended earlier in the deceased group (3 ± 26 and 9 ± 58 min) than the alive group (69 ± 154 and 30 ± 106 min). Mean time for TEVAR was not significantly different in the deceased (28 ± 146 min) versus alive group (35 ± 90 min). Other procedure codes including open chest cardiac massage, cardiopulmonary resuscitation (CPR), and transfusion were associated with poor survival.

**Figure 5.**
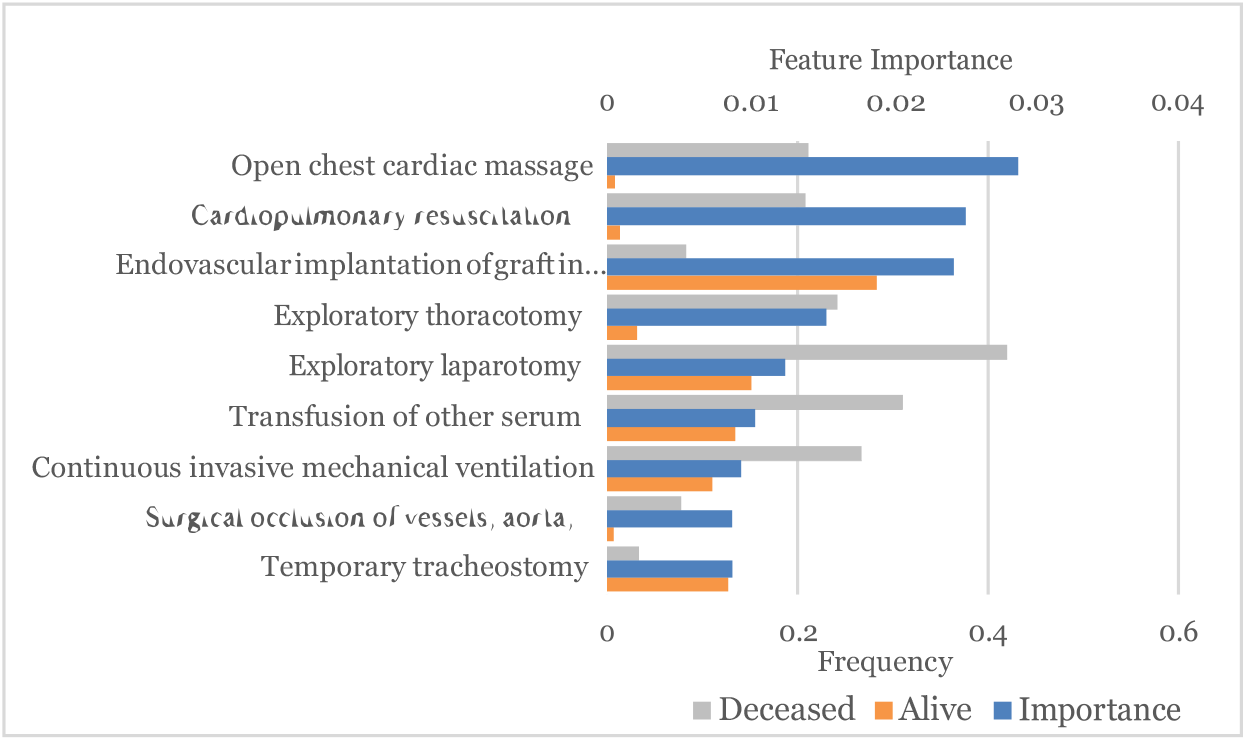
Most predictive procedure codes (PCODE) that contributed to performance of the *Full Data* model. Cardiac massage and CPR were the top predictors for death, while thoracic endovascular graft implantation (TEVAR) was associated with survival. Both exploratory thoracotomy and laparotomy occurred much more frequently in deceased patients.

#### Diagnostic Codes

The highest weighted diagnostic codes in the *Full Data* model were presence of concussion and intracranial hemorrhage, again indicating that head trauma portends a poor prognosis (Figure 6). Splenic injury and abdominal aortic injuries were the next two most important features, indicating that concurrent blunt abdominal injury carries a poor prognosis. Cardiac injury, dissection, and pneumothorax were the remaining top features.

**Figure 6.**
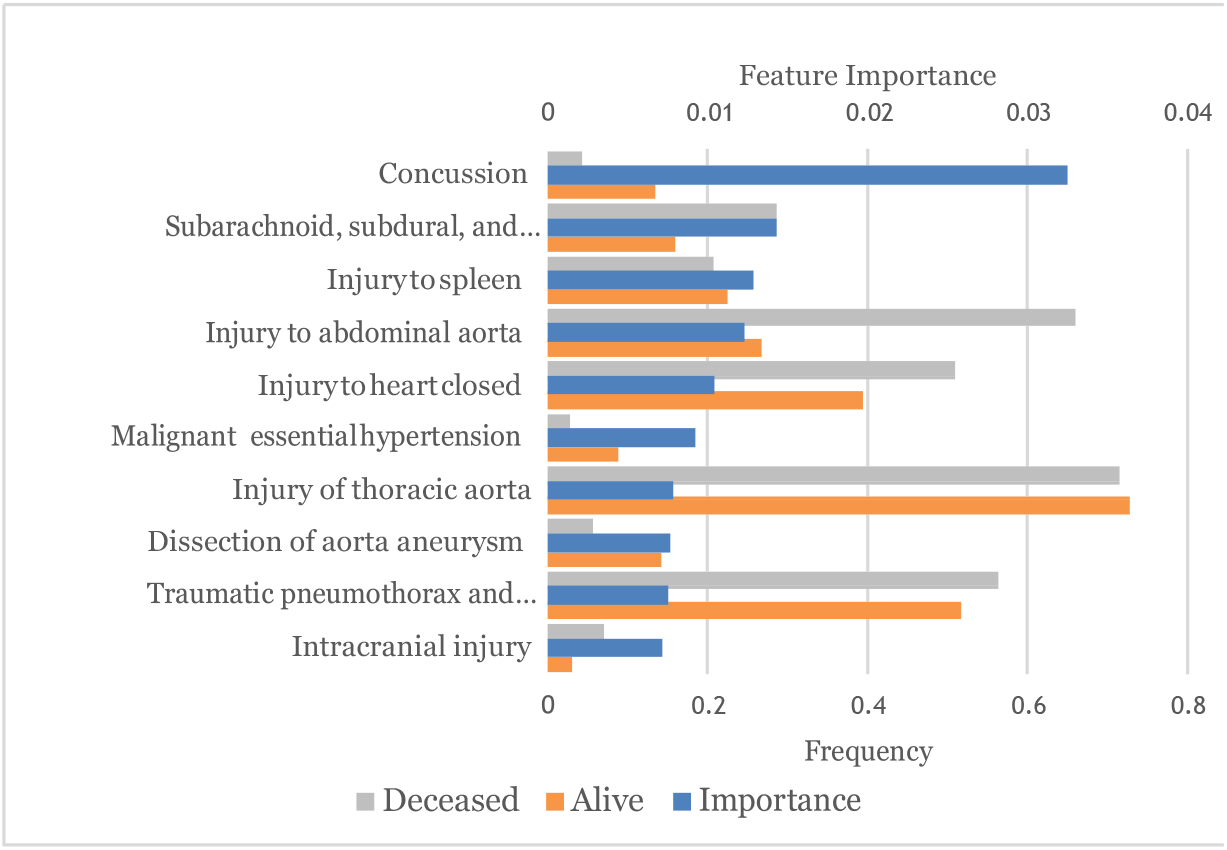
The top 10 most predictive diagnostic codes (DCODE) that contributed to model performance. Head injuries were the two most important diagnostic codes, followed by splenic and abdominal aortic injury indicating blunt abdominal trauma.

#### AIS Codes

Most injury types were more common in the deceased class, including head and facial injuries as previously observed (Figure 7). Abdominal vascular and organ injury were also more common in the deceased group. Interestingly thoracic skeletal and organ injury was slightly more common in the alive group, as was lower extremity injury. This may be related to bias in coding rather than true differences in injury patterns.

**Figure 7.**
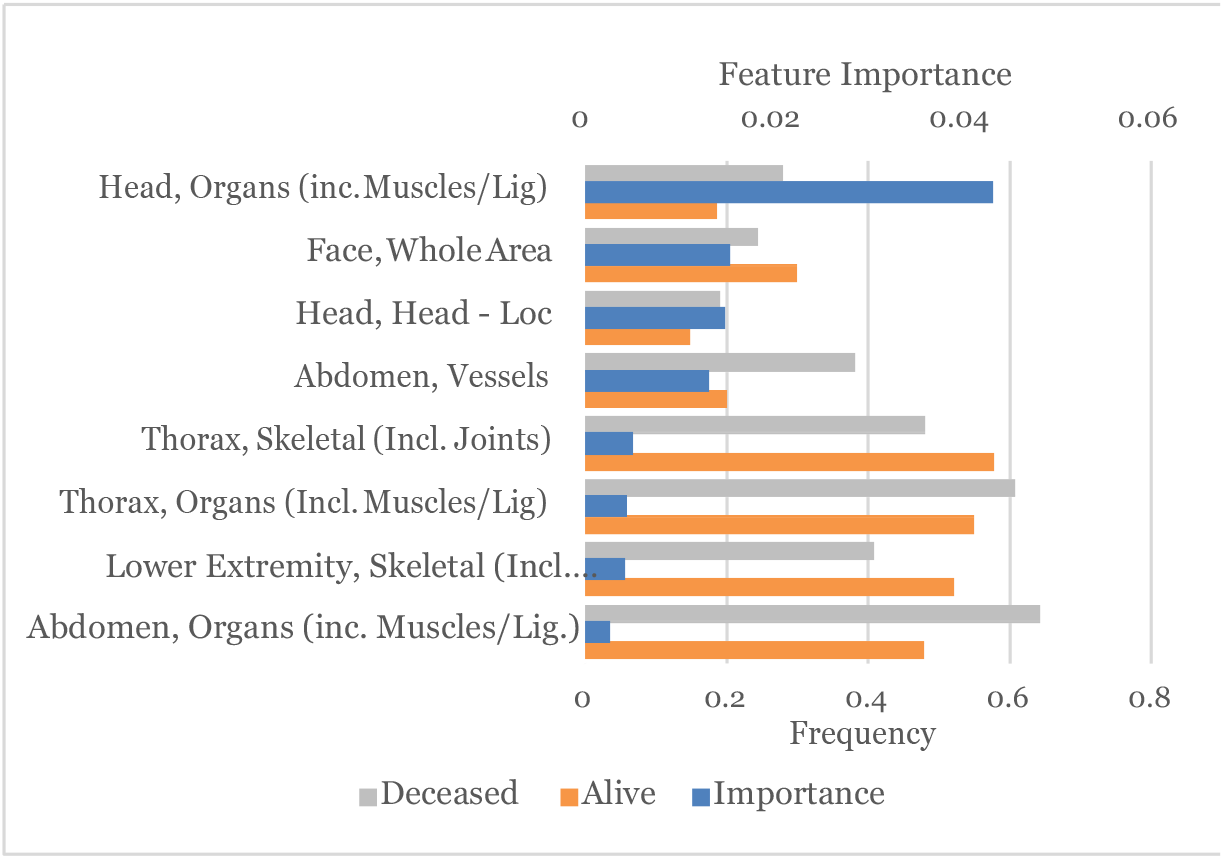
Top predictive AIS codes that contributed to model performance. Head injuries were more common in the deceased class, as were abdominal vascular and organ injury. Concurrent thoracic organ and skeletal injuries were also more common in the deceased class.

**Figure 8.**
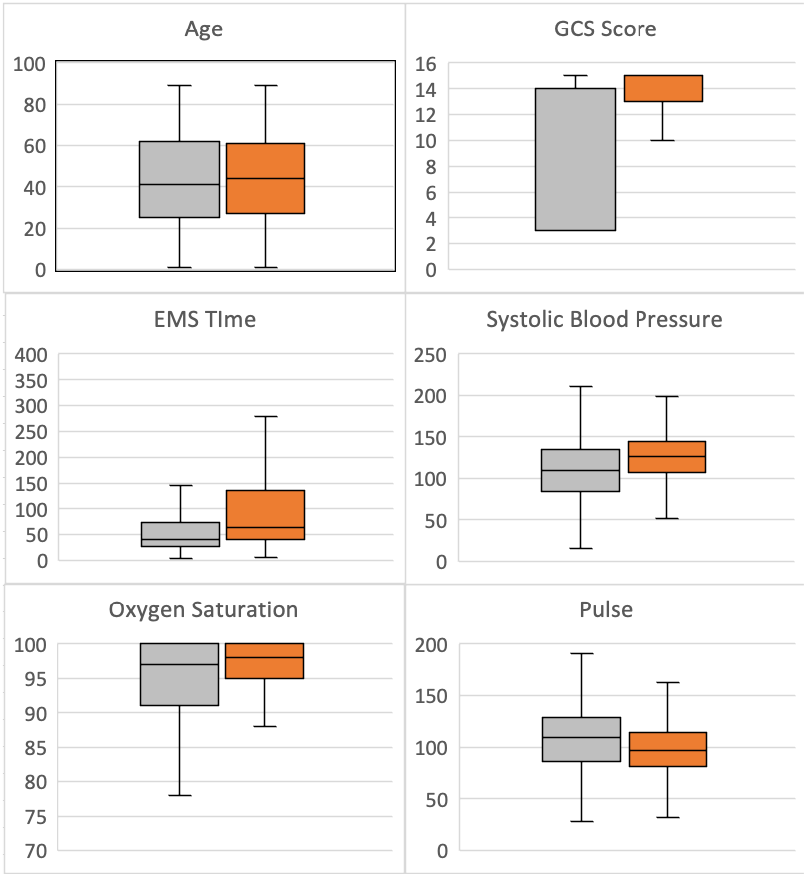
Demographics and vital signs between alive (orange) and deceased (grey) classes. GCS score was lower in the deceased class, whereas other vital signs were similar. Mean EMS time trended lower for the deceased class.

#### Other features

Mean GCS score was 12.6 ± 4.3 in alive patients and 7.5 ± 5.3 in deceased patients (Figure 8). Vital signs including pulse, oxygen saturation, and blood pressure on arrival were not significantly different between classes. Mean time of EMS arrival trended *lower* in the deceased class, suggesting that delay in hospital transfer was not responsible for death in most patients.

## Discussion

We demonstrate that machine learning is a powerful tool to examine factors that contribute to survival in trauma patients. To our knowledge, this is the first published machine learning model for predicting survival in TAI. Our models were able to leverage all available data in the NTDB with no hand-selected features and yield accuracies above 85% in predicting survival. It is interesting to note that the *Full Data* model achieved only slightly higher overall performance that the *Prospective* model despite five out of the top ten features in the *Full Data* model being complications, comorbidities, and diagnostic codes which were not available to the *Prospective* model. This suggests that the *Prospective* model was able to learn substantially different patterns in the data with information available only within the first hour of arrival, and that a patient’s likelihood of survival is largely determined within the first hour of their presentation. However, it is noteworthy that recall for the deceased class suffered the biggest performance drop in the *Prospective* model, indicating that complications or later procedures during hospitalization affect survival. Feature engineering was very important to improving model performance, and our techniques can be re-used by other groups to more easily investigate other clinical questions using the NTDB.

Our results also raise concerns around disparities in outcomes dependent upon race and insurance status. Black patients were more likely to die following TAI, as were poor patients with Medicaid or no insurance (self-pay). Whether this is related to their quality of care cannot be determined, but it is possible that patients of lower socioeconomic status or racial minorities may be more likely to suffer serious injuries, have less access to high quality care, or have worse pre-existing health.

In both models, the GCS score on arrival was the single most important predictor of survival for all patients with TAI. This is expected, as deaths from head injuries account for 34 percent of all traumatic deaths which may overlap with patients with TAI^17^. However, there were no significant differences in age, pulse, or blood pressure on arrival between the classes. Smoking was overall associated with worse increased risk death in both models, and complications of pneumonia and urinary tract infection also contributed to death in the *Full Data* model.

Our results showed that endovascular graft implantation was much more common in patients that survived than those who did not, in keeping with published literature. However, this shows correlation and not necessarily causality because of intrinsic differences in patients who are eligible for graft implantation such as other concurrent injuries, severity of vascular injury, and hemodynamic stability.

Open laparotomy and thoracotomy were highly associated with death in both models and trended earlier in the deceased class, meaning that patients who require these procedures early due to other thoracic or abdominal injuries (e.g. penetrating trauma, bowel injury, etc.) have poor prognosis. Similarly, open chest cardiac massage and CPR were associated with death and it is known that survival following either of these procedures, regardless of root cause, is very low.

This study has limitations. First, model predictions could not be made continuously during an admission due to lack of timestamp data; rather, they could be made shortly after arrival or at the conclusion of care. This means that we could not compare the true hospital course of a patient following specific procedures such as TEVAR. Second, although this dataset is the largest dataset for TAI with 12,435 patients, this is still a relatively small number of samples for traditional machine learning methods and therefore model performance was somewhat lower than more general tools for survival in trauma patients. Lastly, we were unable to externally validate the model on a local dataset due to differences in data structures; however, because the NTDB is sampled from trauma centers nationwide, these results should be generalizable across patient populations and geographies.

In conclusion, we present a machine learning framework for using the NTDB to evaluate factors that contribute to patient survival in patients with traumatic aortic injury, and demonstrate that feature engineering is a crucial step in improving model performance and making results interpretable. All data pre-processing steps and the final model will be released publicly to enable other groups to leverage the NTDB to investigate other important clinical questions.

## Data Availability

We have utilized the National Trauma Data Bank (NTDB) - a large data repository that encompasses a wide variety of traumatic injuries, interventions, and outcomes in trauma patients. The NTDB is compiled annually by the American College of Surgeons (ACS) using standardized data contributions from trauma hospitals across the U.S.

